# Safety and efficiency of the HEART-GP strategy with point-of-care high-sensitivity troponin testing for acute chest pain in out-of-hours primary care: a prospective multicentre diagnostic accuracy study

**DOI:** 10.64898/2026.09.08.26361591

**Authors:** Indra M.B. Melessen, Jelle C.L. Himmelreich, Amy Manten, Eric P. Moll van Charante, Ralf E. Harskamp

## Abstract

**Background:** GPs are often the first clinicians to assess acute chest pain in gatekeeper healthcare systems, but usually with limited diagnostic support. They must distinguish conditions that can be managed safely in primary care from those requiring immediate hospital evaluation, particularly acute coronary syndrome (ACS), while avoiding unnecessary over-referral to already pressured emergency departments. We evaluated HEART-GP, a novel GP-led strategy combining structured clinical assessment, fingerstick point-of-care high-sensitivity troponin I testing, and ECG when available.

**Methods:** We performed a prospective, multicentre, paired diagnostic accuracy study in four Dutch out-of-hours primary care centres implementing the HEART-GP strategy. Patients with acute non-traumatic chest pain and possible ACS according to the treating GP were enrolled from March 2023 to August 2025. Before troponin results were available, GPs recorded intended management, allowing paired comparison with unaided clinical judgement. The HEART-GP strategy recommended urgent referral for high clinical suspicion, troponin of >=4 ng/L, or ischaemic ECG. The primary outcome was adjudicated 6-week major adverse cardiovascular events (MACE). The study was registered as ISRCTN11954040.

**Findings:** Of 1104 patients who underwent HEART-GP, 917 were included. MACE occurred in 62 (6.8%) and myocardial infarction in 52 (5.7%). HEART-GP classified 478 (52.1%) as low risk. For MACE, sensitivity was 95.2% (95% CI 86.5-99.0) and negative predictive value was 99.4% (98.1-99.8); three MACE occurred in the low-risk group, with no cardiac deaths. For myocardial infarction, sensitivity was 98.1% (89.7-100) and negative predictive value was 99.8% (98.6-100). Compared with unaided GP judgement, HEART-GP improved sensitivity for MACE (95.2% vs 67.7%; p<0·001) and myocardial infarction (98.1% vs 67.3%; p<0.001), and reduced urgent referrals (439 [47.9%] vs 483 [52.7%]; p=0.042).

**Interpretation:** HEART-GP showed high rule-out safety and may support GP diagnostic certainty and efficient referrals in patients with possible ACS. Implementation requires training, governance, safety-netting, and local pathways.

**Funding:** Dutch Heart Foundation. Siemens Healthineers provided in-kind device support and had no role in the trial design, conduct or interpretation.

**Research in context:** *Evidence before this study:* Acute chest pain is a common and high-stakes reason for urgent primary care assessment and is the cardinal symptom of acute coronary syndrome. In Dutch out-of-hours primary care, acute chest pain accounts for more than 160,000 contacts annually, or about 9 per 1000 inhabitants. Acute coronary syndrome is ultimately diagnosed in only about 5% of urgent primary care chest pain presentations, but missed cases can be life-threatening; previous primary care studies suggest that 8-19% of ACS cases are not recognised at first assessment. GPs are aware that clinical judgement alone does not reliably meet the very low miss rate considered acceptable for ruling out ACS (around 2–3% in previous work) and therefore often maintain a low threshold for urgent referral despite limited diagnostic support.

*Added value of this study:* This prospective multicentre study evaluated HEART-GP, a GP-led strategy that combines routine clinical assessment with a single fingerstick high-sensitivity cardiac troponin I point-of-care test and ECG findings when available. The HEART-GP strategy showed high rule-out safety, with sensitivity of 95% for major adverse cardiovascular events and 98% for myocardial infarction, and negative predictive values exceeding 99%. Compared with unaided GP assessment, the strategy improved detection of both outcomes while classifying a larger proportion of patients as low risk.

*Implications of all available evidence:* HEART-GP may support safer and more efficient assessment of acute chest pain in out-of-hours primary care by helping GPs identify patients who need immediate hospital evaluation and those who may not require urgent referral. Its observed miss rate approaches the low threshold that GPs have previously indicated as acceptable for ruling out ACS, but the strategy should remain focused on patients in whom ACS is clinically possible but not obvious. Implementation will require training, quality assurance, clear referral pathways, and safety-netting, and further research should assess external validation, cost-effectiveness, patient experience, and effects on ambulance use, emergency department burden, and downstream testing.

## Introduction

Acute chest pain is a common and high-stakes reason for urgent medical contact and the cardinal symptom of acute coronary syndrome (ACS). [1–5] In European gatekeeper healthcare systems, many patients first contact general practitioners (GPs) or out-of-hours primary care (OOH-PC) services rather than ambulance services or emergency departments (EDs). [1,2] In the Netherlands, OOH-PC is delivered by regional GP cooperatives and records more than 160,000 acute chest pain contacts annually, equivalent to about 9 contacts per 1000 inhabitants. [3]

The central challenge for GPs is to decide, with limited diagnostic support, which patients with acute chest pain require urgent hospital assessment and which can be managed safely without immediate referral. The most feared diagnosis is ACS: although present in only about 5% of urgent primary-care chest pain presentations, delayed recognition can forfeit the opportunity for timely treatment and lead to preventable myocardial injury, serious complications, or death. [3–7] Guidelines therefore recommend a low referral threshold when ACS is suspected. [6] Although this safety-first approach is appropriate, it is both inefficient and incomplete: more than half of patients are referred despite most not having serious cardiac disease, while clinical suspicion alone still misses 8% to 19% of ACS cases in primary care. [3,5,7,8]

Emergency department pathways address this diagnostic uncertainty by combining structured risk assessment with high-sensitivity cardiac troponin testing. [4] However, these pathways depend on infrastructure that is often unavailable in out-of-hours primary care, including rapid laboratory access, observation capacity, and serial testing. Earlier primary-care studies using non-high-sensitivity troponin tests lacked sufficient rule-out safety. [5] More recent evidence suggests that high-sensitivity troponin can support rule-out outside hospital when rapid laboratory testing is available, and that single-sample fingerstick point-of-care high-sensitivity cardiac troponin I testing can safely rule out myocardial infarction in low-risk emergency department patients. [9,10]

Whether such a single-sample point-of-care strategy can safely bridge the gap to out-of-hours primary care remained uncertain. We therefore developed HEART-GP as a pragmatic GP-led risk stratification strategy for acute non-traumatic chest pain in this setting. HEART-GP combines routine GP assessment, a single fingerstick high-sensitivity cardiac troponin I point-of-care test, and ECG findings when available. [12] Its intended clinical role is to support referral decisions in the diagnostic grey zone: patients in whom ACS is possible but not already clinically obvious. In this study, we aimed to evaluate the diagnostic safety and efficiency of HEART-GP for ruling out 6-week ACS and other major adverse cardiovascular events (MACE), and to compare HEART-GP with unaided GP clinical judgement recorded before troponin results were available.

## Methods

### Study design and setting

We performed a prospective, multicentre, paired diagnostic accuracy study in four Dutch OOH-PCs located in Amersfoort, Alkmaar, Leiderdorp, and Venlo. These centres are situated across four geographically distinct regions of the Netherlands (central, north-western, western, and southern) and serve both urban and more rural populations. Together, they cover a catchment population of approximately one million inhabitants, corresponding to about 5-6% of the Dutch population. Dutch OOH-PCs are regional GP cooperatives staffed by GPs and supported by trained triage nurses. After telephone triage, patients may receive telephone advice, be invited for face-to-face GP consultation at the centre, receive a home visit, or be referred directly to ambulance or hospital care. In this study, recruitment took place among patients assessed by the treating GP for acute chest pain in whom ACS was considered a possible diagnosis. Study coordination was centralised at the Department of General Practice of Amsterdam UMC, which managed consent procedures and follow-up with participants, GPs, and hospitals. Eligible patients provided initial verbal consent during the acute consultation, followed by written informed consent coordinated centrally by the study team; no separate informed consent was obtained from treating GPs. The study was prospectively registered in the Dutch national OMON/ToetsingOnline (NL-OMON51574) and international ISRCTN registry (ISRCTN11954040) and approved by the Medical Ethics Committee of Amsterdam UMC (NL82428.000.22), after initial review by the national medical ethics committee (CCMO). This report follows STARD guidance for diagnostic accuracy studies and is intended for reporting alongside a STARD checklist. [11] The protocol and detailed methods have been published previously. [12]

### Participants

Consecutive adults aged 18 years or older were eligible if they presented with acute non-traumatic chest pain and the treating GP considered ACS a possible diagnosis. Patients were enrolled between March 17, 2023, and Aug 1, 2025. The intended-use population comprised patients in whom ACS was clinically plausible but not already evident. Patients with a very high a priori likelihood of ACS, such as ST-segment elevation on ECG or a presentation warranting immediate referral, were not the intended target population. Patients in whom ACS was not considered clinically plausible were also outside the intended-use population. Patients were excluded from the analysis if they were haemodynamically unstable, unable to provide final written informed consent, or not registered with a Dutch GP, which was required for follow-up. Patients with recurrent presentations during enrolment were eligible only at first presentation.

### Index strategy

The HEART-GP strategy, as illustrated in ***Figure 1***, was applied during the index consultation. GPs completed a brief structured checklist covering chest pain characteristics, associated symptoms, whether the presentation was compatible with ACS, and the degree of clinical concern, including gut feeling. A single high-sensitivity cardiac troponin I point-of-care measurement was done using fingerstick sampling with the Siemens Atellica VTLi assay. ECG assessment was included when available. The prespecified point-of-care troponin rule-out threshold was below 4 ng/L, based on previous diagnostic safety data for this assay in low-risk patients and well below the 99th percentile upper reference limit of 23 ng/L. [10,13] Urgent referral was recommended if there was high clinical suspicion, point-of-care high-sensitivity troponin I of >=4 ng/L, or ECG abnormalities suggestive of myocardial ischaemia. When all these criteria were absent, no urgent referral was recommended. For symptoms lasting less than two hours, GPs were instructed not to use troponin or ECG results for rule-out. If troponin testing failed or uncertainty remained, management relied on clinical judgement, with guideline consultation or cardiologist advice when needed. The final management decision remained at the discretion of the treating GP. Before the troponin result became available, GPs recorded intended management based on clinical assessment alone. Once submitted, this intended decision could not be revised. This procedure enabled paired comparison of HEART-GP with unaided GP judgement within the same patients.

**Figure 1.**
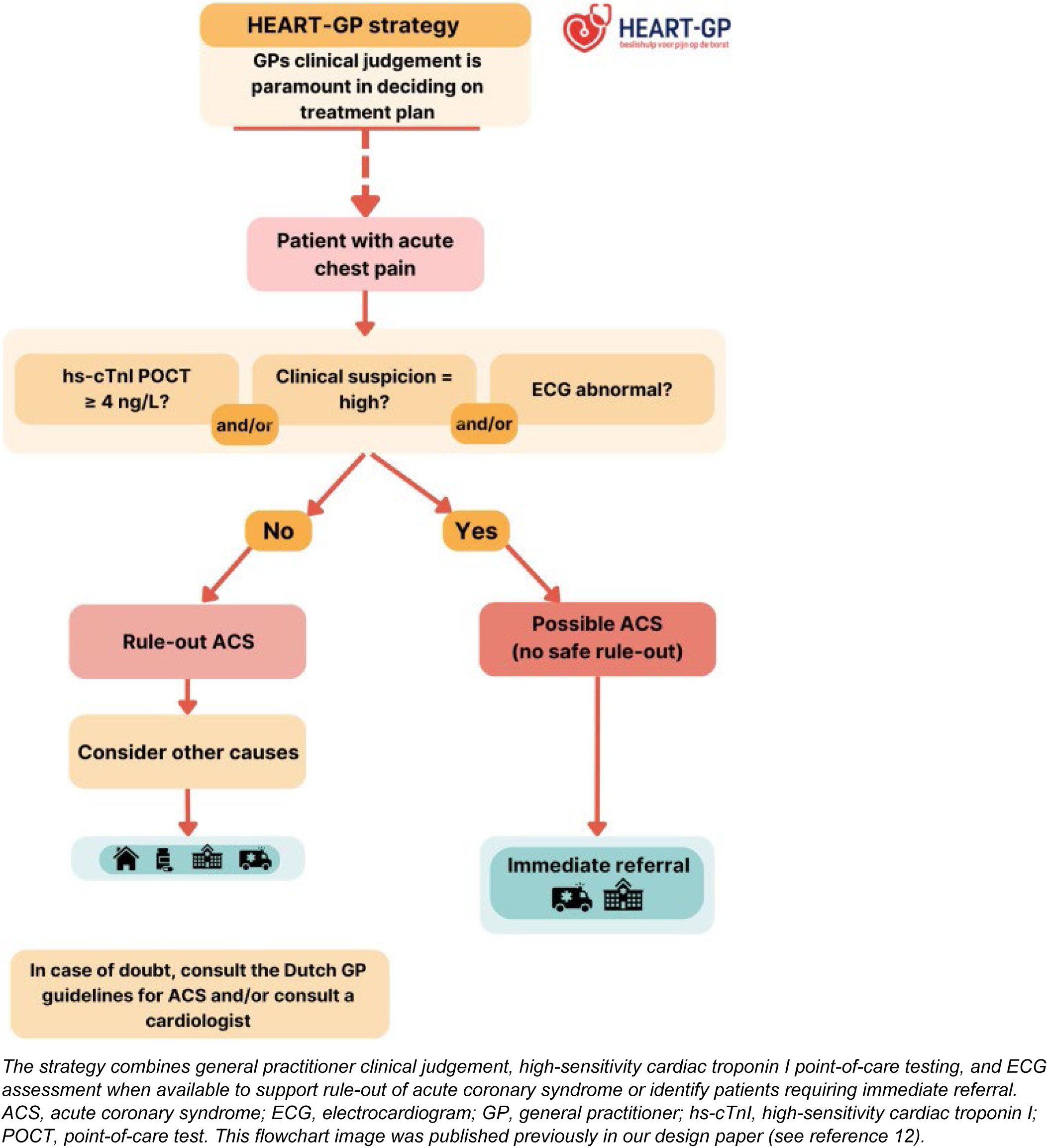
HEART-GP risk stratification strategy for patients with acute chest pain in out-of-hours primary care

### Reference standard and outcomes

We followed participants for 6 weeks after the index consultation. Patients were contacted by telephone for self-reported outcomes, and follow-up information was requested from their GP, and ED and specialist correspondence was obtained from hospitals when relevant. In the Dutch healthcare system, the GP medical record serves as the central longitudinal record for patient care and includes information on hospital admissions, diagnostic testing, procedures, and specialist follow-up. The primary outcome was 6-week MACE, defined a priori as all-cause mortality, ACS, or urgent coronary revascularisation. ACS included ST-elevation myocardial infarction (STEMI), non-ST-elevation myocardial infarction (NSTEMI), and unstable angina. The secondary safety outcome was 6-week myocardial infarction (MI), defined as STEMI or NSTEMI. Clinical outcomes were adjudicated by a panel of three clinicians (two GPs, one cardiologist) blinded to point-of-care troponin results. We assessed safety by sensitivity and NPV for MACE and MI. Efficiency was our co-secondary outcome, defined as the proportion of patients classified as low risk/no urgent referral.

### Patient and stakeholder involvement

Patient, public, and professional involvement informed the development and implementation of HEART-GP. The study was discussed with the Dutch Heart Foundation, a patient panel, executive board members, and health-care professionals from participating out-of-hours primary care cooperatives. Patients were also contacted personally to share experiences with the strategy and provide feedback on risk communication; these findings will be reported separately. Study findings will be disseminated through participating cooperatives and Dutch Heart Foundation networks.

### Sample size and statistical analysis

The minimum sample size was 845 patients to estimate sensitivity with sufficient precision, assuming a 6-week MACE rate of approximately 7%; our target sample size was set at least 900 included patients [12]. We assessed diagnostic accuracy by calculating sensitivity, specificity, positive predictive value (PPV), and negative predictive value (NPV) with corresponding 95% confidence intervals (95% CIs). Efficiency was calculated as the proportion of patients not urgently referred, including both true-negative and false-negative classifications. Missing baseline covariates were handled using multiple imputation by chained equations with five imputed datasets, under a missing-at-random assumption. Analyses were done using SPSS software (IBM Corp. IBM SPSS Statistics for Windows. Version 31 Armonk, NY: IBM Corp; 2025). Categorical variables are reported as numbers and percentages, and continuous variables as medians and IQRs. For paired comparisons between HEART-GP and unaided GP judgement, absolute paired differences in proportions were calculated from discordant cells within each imputed dataset and pooled using Rubin’s rules, yielding pooled differences, 95% CIs, and p values.

## Results

### Patient flow and baseline characteristics

Between March 17, 2023, and Aug 1, 2025, 1104 patients underwent the HEART-GP strategy based on verbal consent. Of these, 917 provided written informed consent, met final inclusion criteria, and had complete 6-week follow-up for MACE; these participants were included in the diagnostic accuracy analysis (also see ***Figure 2***). Detailed clinical data could not be collected for the 187 patients who did not provide full written consent, but comparison of limited available baseline information did not indicate major differences in age, sex, symptom duration, or troponin test results.

**Figure 2.**
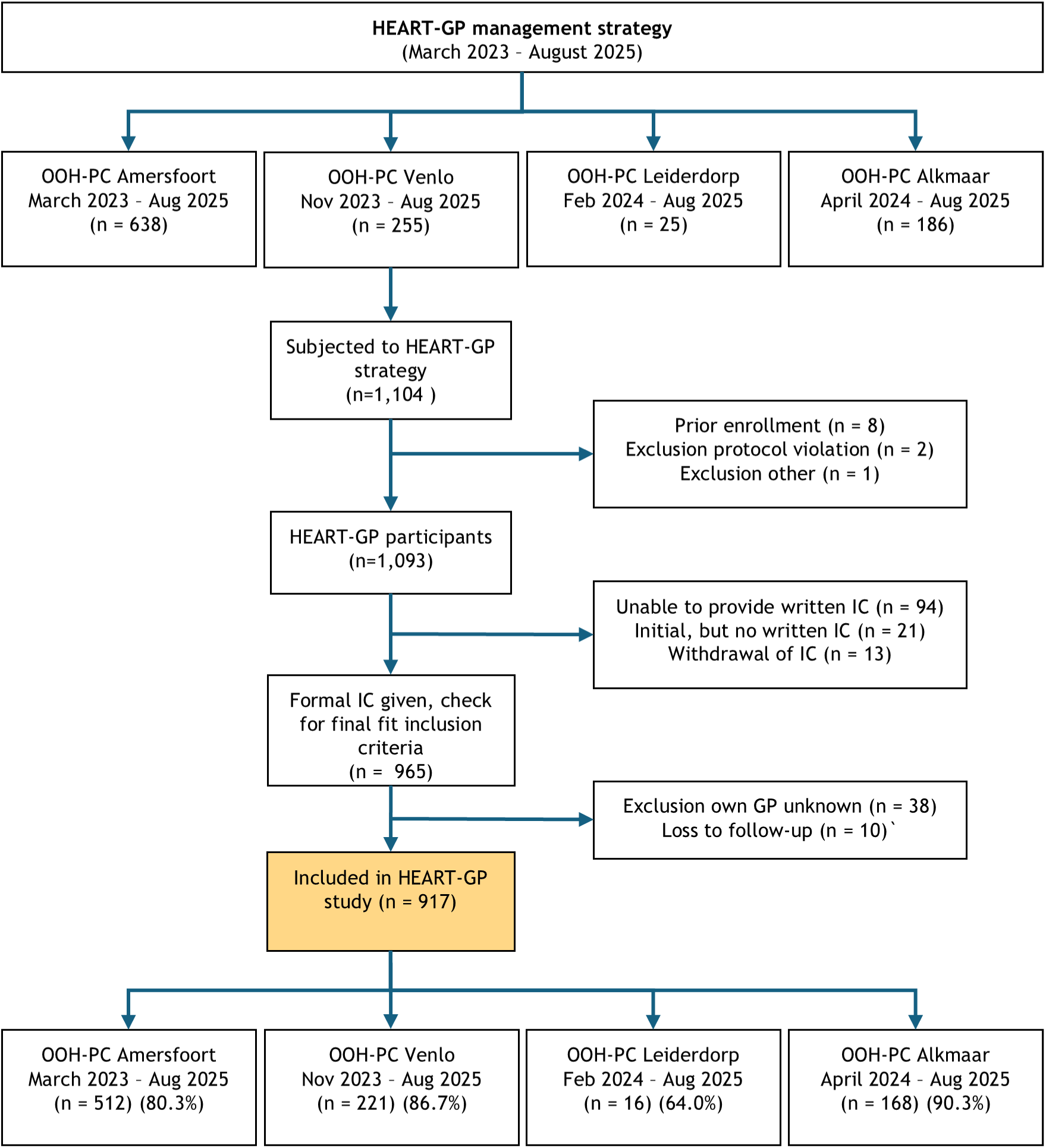
Flowchart of recruitment per site and exclusions

Baseline characteristics are presented in ***Table 1***. The median age was 54 years (IQR 41-66), and 483 (52.7%) participants were female. Most participants had at least one cardiovascular risk factor (707 [77.1%]). Chest pain presentations were heterogeneous: pressing chest pain was reported by 677 (73.8%) participants, radiation to the arm by 352 (38.4%), nausea or sweating by 386 (42.1%), and dyspnoea by 403 (43.9%). Median symptom duration before presentation was 12 h (IQR 3.5-24).

**Table 1.** Baseline characteristics of the study population.

| Characteristic | All patients<br>(n = 917) | MACE<br>(n = 62) | No MACE<br>(n = 855) |
| --- | --- | --- | --- |
| Female sex, n (%) | 483 (52.7) | 22 (35.5) | 461 (53.9) |
| Age, years, median (IQR) | 54 (41 to 66) | 66 (54 to 72) | 54 (40 to 65) |
| Ethnicity/cultural background<sup>1</sup> |  |  |  |
| Dutch | 745 (81.2) | 54 (87.1) | 691 (80.8) |
| Western | 79 (8.6) | 4 (6.5) | 75 (8.8) |
| Non-Western | 96 (10.5) | 2 (3.2) | 94 (11.0) |
| Other | 26 (2.8) | 0 (0) | 26 (3.0) |
| **Risk factor for CVD, n (%)** |  |  |  |
| Any risk factor, listed below | 707 (77.1) | 54 (87.1) | 653 (76.4) |
| Current smoker | 190 (20.7) | 13 (21.0) | 177 (20.7) |
| Missing, n (%) | 31 (3.4) | 2 (3.2) | 29 (3.4) |
| BMI >30 | 228 (24.9) | 19 (30.6) | 209 (24.4) |
| Missing, n (%) | 50 (5.5) | 2 (3.2) | 48 (5.6) |
| Family history of CVD | 394 (43.0) | 30 (48.4) | 364 (42.6) |
| Missing, n (%) | 57 (6.2) | 6 (9.7) | 51 (6.0) |
| Hypertension | 307 (33.5) | 35 (56.5) | 272 (31.8) |
| Missing, n (%) | 29 (3.2) | 1 (1.6) | 28 (3.3) |
| Hypercholesterolaemia | 261 (28.5) | 40 (64.5) | 221 (25.8) |
| Missing, n (%) | 50 (5.5) | 2 (3.2) | 48 (5.6) |
| Diabetes mellitus | 82 (8.9) | 11 (17.7) | 71 (8.3) |
| Missing, n (%) | 28 (3.1) | 0 (0) | 28 (3.3) |
| Prior myocardial infarction | 108 (11.8) | 16 (25.8) | 92 (10.7) |
| Prior PCI or CABG | 112 (12.2) | 16 (25.8) | 96 (11.2) |
| Prior CVA or TIA | 33 (3.6) | 3 (4.8) | 30 (3.5) |
| Prior peripheral arterial disease | 18 (2.0) | 1 (1.6) | 17 (2.0) |
| **Presenting symptoms, n (%)<sup>1</sup>** |  |  |  |
| Symptom duration, hours, median (IQR) | 12 (3.5 to 24) | 12 (4 to 24) | 12 (3.5 to 24) |
| Missing, n (%) | 16 (1.7) | 2 (3.2) | 14 (1.6) |
| Pressing type of chest pain | 677 (73.8) | 48 (77.4) | 629 (73.6) |
| Radiation to arm | 352 (38.4) | 27 (43.5) | 325 (38.0) |
| Nausea and/or sweating | 386 (42.1) | 25 (40.3) | 361 (42.2) |
| Increase with exercise | 301 (32.8) | 25 (40.3) | 276 (32.3) |
| Dyspnoea | 403 (43.9) | 25 (40.3) | 378 (44.2) |
| Chest pain with palpation | 247 (26.9) | 8 (12.9) | 239 (28.0) |

Abbreviations: BMI, body mass index; CABG, coronary artery bypass grafting; CVA, cerebrovascular accident; CVD, cardiovascular disease; GP, general practitioner; hs, high-sensitivity; IQR, interquartile range; OOH-PC, out-of-hours primary care; PCI, percutaneous coronary intervention; POCT, point-of-care test; TIA, transient ischaemic attack. In cases of missingness, number (%) of missing per variable are shown per variable. If missingness was not reported in the table, the variable had complete data.
<sup>1</sup> Response categories not mutually exclusive, totals could add up to over 100%.

### Clinical management after HEART-GP assessment

After HEART-GP assessment, 439 (47.9%) participants were referred for urgent care and 478 (52.1%) were classified as low risk without immediate referral. Among referred participants, 54 (16%) were referred because of persistent clinical concern or ischaemic ECG changes despite a troponin result below the prespecified 4 ng/L rule-out threshold. Among non-referred participants, 101 (21%) had a troponin concentration at or above 4 ng/L, all below the 99th percentile upper reference limit of 23 ng/L. Clinical notes indicated that GPs interpreted these low-level troponin elevations in the context of the full presentation, including symptoms, estimated likelihood of ACS, clinical concern or gut feeling, and ECG findings. Troponin testing failed or yielded no result in 40 (4.4%) participants; in these cases, GPs made management decisions without a troponin result. (***Supplementary file, Table S1***).

### Clinical outcomes

Within six weeks, 62 (6.8%) participants experienced MACE. These events included two deaths, 15 STEMIs, 37 NSTEMIs, eight unstable angina events, and one urgent revascularisation for rapidly progressive angina outside an adjudicated ACS event. The two deaths comprised one cardiac death after STEMI and one non-cardiac death due to cholangiocarcinoma. Overall, 52 (5.7%) participants had myocardial infarction.

### Diagnostic outcomes

The HEART-GP strategy identified 59 of 62 MACE events, yielding sensitivity of 95.2% (95% CI 86.5 to 99.0) and NPV of 99.4% (95% CI 98.1 to 99.8) (***Table 2***). Specificity was 55.6% (95% CI 52.2 to 58.9), and PPV was 13.4% (95% CI 12.4 to 14.6). No participant classified as low risk died of a cardiac cause. The three MACE events among low-risk participants comprised one MI, one unstable angina event, and one non-cardiac death (***Supplementary file, Table S2***).

**Table 2.** Diagnostic accuracy of HEART-GP for 6-week MACE and myocardial infarction (N = 917)

| **Outcome** | **Events** | **TP** | **FN** | **FP** | **TN** | **Sensitivity** | **Specificity** | **PPV** | **NPV** |
| --- | --- | --- | --- | --- | --- | --- | --- | --- | --- |
| **MACE** | 62<br>(6.8%) | 59 | 3 | 380 | 475 | 95.2%<br>(86.5-99.0) | 55.6%<br>(52.2-58.9) | 13.4%<br>(12.4-14.6) | 99.4%<br>(98.1-99.8) |
| **Myocardial infarction** | 52<br>(5.7%) | 51 | 1 | 388 | 477 | 98.1%<br>(89.7-100) | 55.1%<br>(51.8-58.5) | 11.6%<br>(10.8-12.5) | 99.8%<br>(98.6-100) |

Values are n unless otherwise indicated. Events are presented as n/N (%). Diagnostic accuracy estimates are presented as % (95% CI). FN, false-negative; FP, false-positive; HEART-GP, the HEART-GP strategy; MACE, major adverse cardiovascular events; NPV, negative predictive value; PPV, positive predictive value; TN, true-negative; TP, true-positive; 95% CI, 95% confidence interval.

For myocardial infarction, HEART-GP identified 51 of 52 events, corresponding to sensitivity of 98.1% (95% CI 89.7 to 100) and NPV of 99.8% (95% CI 98.8 to 100). Specificity was 55.1% (95% CI 51.8 to 58.5) and PPV was 11.6% (95% CI 10.8 to 12.5). The initially missed myocardial infarction occurred in a participant who presented with chest pain, a normal ECG, low point-of-care troponin, and symptom resolution after nitroglycerin after a total symptom duration of less than 15 minutes. Chest pain recurred later the same evening, anterior STEMI was diagnosed, and primary PCI of the left anterior descending artery was performed. The participant recovered without complications and was discharged in good condition three days later.

### Comparison with unaided GP judgement

Unaided GP judgement identified 42 of 62 MACE events and missed 20, corresponding to sensitivity of 67.7% (95% CI 54.7-79.1) and negative predictive value of 95.4% (93.5-96.8). HEART-GP classified 18 of these 20 initially missed MACE events for urgent referral, while one MACE event that would have been referred according to unaided judgement was classified as non-urgent. Paired analyses, as illustrated in ***Figure 3***, showed higher sensitivity and negative predictive value for MACE with HEART-GP than with unaided judgement (p<0·001 for both comparisons).

**Figure 3.**
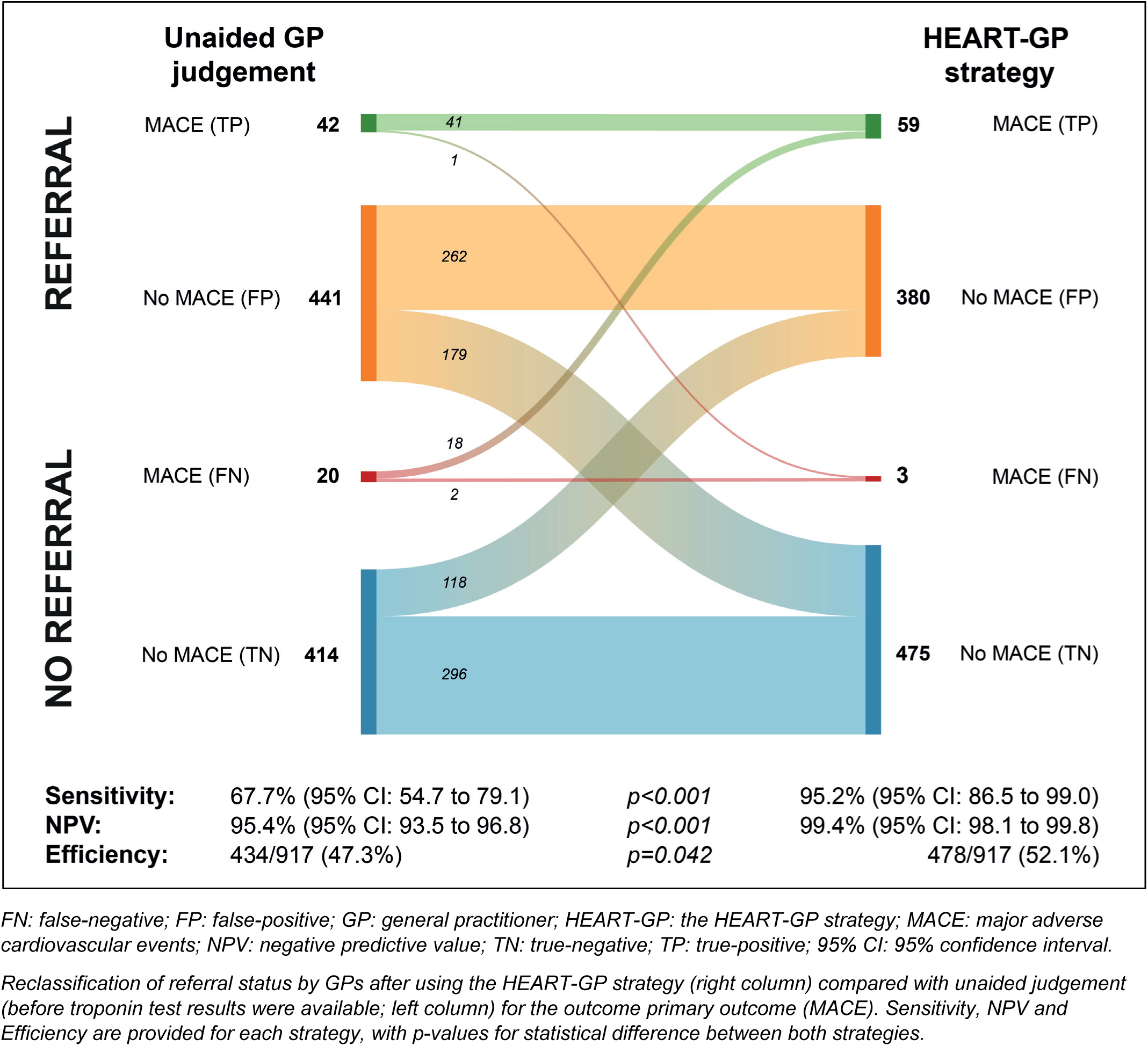
Comparison of HEART-GP with unaided GP judgement for MACE.

For myocardial infarction, unaided GP judgement identified 35 of 52 events and missed 17, yielding sensitivity of 67.3% (95% CI 52.9-79.7) and negative predictive value of 96.1% (94.3-97.3). HEART-GP classified all 17 myocardial infarctions missed by unaided judgement for urgent referral, while one myocardial infarction initially selected for referral by unaided judgement was classified as non-urgent (***Supplementary file, Figure S1***). Sensitivity and negative predictive value for myocardial infarction were higher with HEART-GP than with unaided GP judgement (p<0·001 for both comparisons).

HEART-GP also modestly reduced urgent referrals. Unaided GP judgement would have resulted in urgent referral of 483 (52.7%) participants, compared with 439 (47.9%) after HEART-GP, an absolute reduction of 4.8 percentage points (95% CI 0.2-9.4; p=0·042).

## Discussion

### Principal findings

In this prospective multicentre diagnostic accuracy study in Dutch OOH-PC, HEART-GP showed high rule-out safety for 6-week MACE and myocardial infarction among selected patients with acute non-traumatic chest pain and possible ACS. No patient classified as low risk died of a cardiac cause during follow-up. Compared with unaided GP judgement recorded before troponin results were available, HEART-GP identified more patients who subsequently had MACE or myocardial infarction and modestly reduced urgent referrals. These findings suggest that adding point-of-care high-sensitivity troponin testing to structured GP assessment can support safer and more efficient referral decisions in the intended-use population.

### Strengths and limitations

Several limitations should be considered. The study population was defined by patients with acute chest pain in whom the treating GP considered ACS a possible diagnosis. This reflects the intended-use population of HEART-GP, but it also means that not all chest pain presentations were eligible or included; patients with an obvious non-cardiac explanation, such as clear costomyalgia, would generally not have been approached. Because this initial clinical judgement was part of routine care, we could not verify eligibility among all chest pain contacts, and selection bias cannot be excluded. Nevertheless, inclusion of eligible patients proceeded well, with high recruitment across participating centres, supporting the feasibility of embedding HEART-GP into routine out-of-hours primary care. The number of outcome events limited the precision of sensitivity estimates and precluded robust subgroup analyses. Because point-of-care troponin results informed clinical management, GPs could not be blinded. This study therefore evaluates HEART-GP as a clinical strategy, not the diagnostic performance of the troponin assay in isolation. The comparison with unaided GP assessment was based on intended management decisions recorded on a paper checklist before point-of-care troponin results were available. Once completed, these forms could not be altered in response to the test result, reducing the risk that the pre-test decision was retrospectively changed. In addition, clinical records frequently documented changes in management after HEART-GP results became available, supporting the validity of the paired comparison. Finally, although 6-week MACE as a delayed reference standard reduced the risk of partial verification bias, it cannot eliminate it completely.

### Comparison with other studies

These findings are consistent with the central role of high-sensitivity cardiac troponin in hospital-based acute chest pain pathways [4]. However, ED pathways cannot simply be transferred to OOH-PC. Primary care patients are less selected, disease prevalence is lower, serial testing and observation are often not feasible, and ECGs or laboratory troponin results may not be routinely available. The recent ESC scientific statement on point-of-care high-sensitivity troponin emphasises that such assays should be evaluated with the intended users and in the intended clinical setting [14].

The Norwegian OUT-ACS implementation study showed that a 0/1-hour laboratory-based high-sensitivity troponin strategy could be implemented in emergency primary care and was associated with rapid rule-out of MI outside hospital [9]. The subsequent OUT-POC pilot study suggested that high-sensitivity troponin point-of-care testing in emergency primary care is feasible when used by non-laboratory personnel, but it was small and primarily designed as a pilot evaluation [15]. HEART-GP study adds to this evidence by evaluating a pragmatic single-consultation strategy in four Dutch OOH-PC populations. Rather than testing troponin as an isolated diagnostic test, HEART-GP evaluates the clinical role most relevant to GPs: whether adding immediate high-sensitivity troponin point-of-care testing to routine assessment improves the safety of referral decisions.

Related Dutch primary care research, including the POB-HELP trial, evaluates rule-out strategies in daytime primary care, a setting with a lower-risk and different workflow than OOH-PC [16]. HEART-GP therefore addresses a complementary but distinct urgent-care population in which decisions about ambulance transport and urgent hospital referral are common.

Unaided clinical judgement showed a sensitivity of 65–70%, which is consistent with previous Dutch OOH-PC studies reporting sensitivities well below 90% [17–19]. This performance remains below what is generally considered acceptable for safely ruling out ACS. In a national survey among Dutch GPs, clinicians acknowledged the fallibility of clinical judgement in acute chest pain and expressed strong support for additional diagnostic support, including clinical decision aids, troponin point-of-care testing, or a combination of both [20]. More recently, Johannessen *et al* argued that decision tools in emergency primary care should strive for a miss rate of only 2-3%, corresponding to a sensitivity of 97–98%, a threshold that is supported by earlier ED literature using pretest probability thresholds below 2% and by a systematic review in which clinical opinion indicated a minimum acceptable sensitivity of 97% [21–23]. In this context, the HEART-GP strategy addresses a clear clinical need and meets the predefined practitioner requirements for both safety and efficiency.

### Implications for practice and further research

Although further external validation is warranted, the potential relevance of HEART-GP may extend beyond the Dutch gatekeeper system to other urgent primary care settings where immediate laboratory-based troponin testing is unavailable. HEART-GP may help GPs move beyond high-risk symptoms and clinical gestalt alone by adding an objective biomarker result to routine assessment. However, the strategy should be understood as a safety-focused rule-out tool for the diagnostic grey zone: patients in whom ACS is considered possible but not clinically obvious. This grey zone represents a substantial proportion of chest pain presentations, but requires careful clinical judgement. HEART-GP should not be applied when ACS is not deemed clinically relevant, as indiscriminate troponin testing may lead to diagnostic creep, false-positive findings, downstream testing, and unnecessary referral. Conversely, in clear-cut ACS or clinically unstable patients, HEART-GP should not delay immediate referral or emergency care. In patients with reassuring clinical assessment and troponin below the predefined threshold, HEART-GP can support non-referral with clear safety-netting advice. In patients with elevated troponin, test failure, persistent or progressive symptoms, abnormal clinical findings, or ongoing GP concern, referral or specialist consultation remains appropriate.

Successful implementation requires more than availability of a point-of-care device. In our study, GPs and assistants received dedicated training, supported by practical guides and instructional videos, on when and how to use the HEART-GP strategy. Such training is essential to prevent inappropriate use outside the intended target population. Implementation also requires clear device-specific protocols, quality assurance supported by clinical chemistry, and agreed pathways with ambulance services, emergency departments, and cardiology. Because point-of-care troponin values may be confused with hospital laboratory troponin results, communication should explicitly state which device was used and which cut-off applies. Reporting results in predefined risk categories alongside the numerical value may further reduce misinterpretation.

Further research should evaluate cost-effectiveness, patient experience, implementation barriers, and the effect of HEART-GP on ambulance use, ED crowding, and downstream testing. Additional work should examine whether incorporating ECG, symptom duration, serial point-of-care testing in selected patients, or alternative high-sensitivity point-of-care assays can further improve safety without undermining feasibility in OOH-PC.

## Conclusion

In patients presenting to OOH-PC with acute non-traumatic chest pain, the HEART-GP strategy demonstrated high rule-out safety for MACE and MI and outperformed unaided GP assessment. By integrating high-sensitivity troponin point-of-care testing into routine GP assessment, HEART-GP improved rule-out safety while classifying a higher proportion of patients as low risk. These findings support HEART-GP as a practical and safe risk stratification tool to guide referral decisions in low-risk, high-volume urgent primary care settings.

## Supporting information

STARD checklist

## Contributors and guarantor

IMBM coordinated the day-to-day conduct of the study, performed the main data analyses, and drafted the initial version of the manuscript. JCLH contributed substantially to the data analyses, interpretation of findings, and manuscript writing. AM contributed to the interpretation of findings, and manuscript editing. EPMvC contributed to the study methodology, interpretation of findings, manuscript editing, and overall supervision. REH conceived the study, obtained funding, supervised the project, and led manuscript writing and revision. All authors reviewed and approved the final manuscript. REH is the guarantor and accepts full responsibility for the conduct of the study, had access to the data, and controlled the decision to publish.

## Acknowledgements

The authors thank Edanur Sert (research coordinator, Amsterdam UMC), Simone van den Bulk (general practitioner, Leiden University Medical Center), Tobias Bonten (general practitioner, Leiden University Medical Center), Martijn H. Rutten (general practitioner, Radboud university medical center and OOH-PC Venlo), Pier Woudstra (interventional cardiologist, Frisius Medical Center), Wim Lucassen (general practitioner, Zwaag), and Wim Busschers (statistician, Amsterdam UMC) for their contributions to the study. We also thank the participating OOH-PC centres (Huisartsenpost Eemland, HONK, Cohesie, and LIMES) and their staff, including Jack Baaij, Tirza Volk, Carla van Velden-Hollander, Natalie Fermont-Fleuren, Saine Sinnecker, Remco Rietveld, Corrie Vellema, and Liesbeth van der Plas. Finally, we thank all patients who participated in the HEART-GP study.

## Data sharing

Individual participant data are not publicly available because participant consent and data governance arrangements allow use of the study data within Amsterdam UMC and collaborating research arrangements only. Researchers interested in additional analyses or collaboration can contact the corresponding author or submit a request via www.amstelheart.nl. Requests will be considered by the study team and, where appropriate, analyses may be performed in collaboration with external investigators, subject to scientific relevance, ethical approval where required, and applicable Dutch and European data protection regulations. Aggregated data supporting the findings of this study can be shared upon reasonable request.

## AI declaration

OpenAI’s ChatGPT was used to support grammar, spelling, wording, and editorial refinement during manuscript preparation. The tool was not used to generate, analyse, or interpret study data. All AI-assisted text was reviewed, edited, and verified by the authors, who take full responsibility for the accuracy, integrity, and final content of the manuscript.

