## Supplemental data file for "Safety and efficiency of the HEART-GP strategy with point-of-care high-sensitivity troponin testing for acute chest pain in out-of-hours primary care: a prospective multicentre diagnostic accuracy study"

**Supplementary Figure S1. Comparison of HEART-GP with unaided GP judgement for MI.**

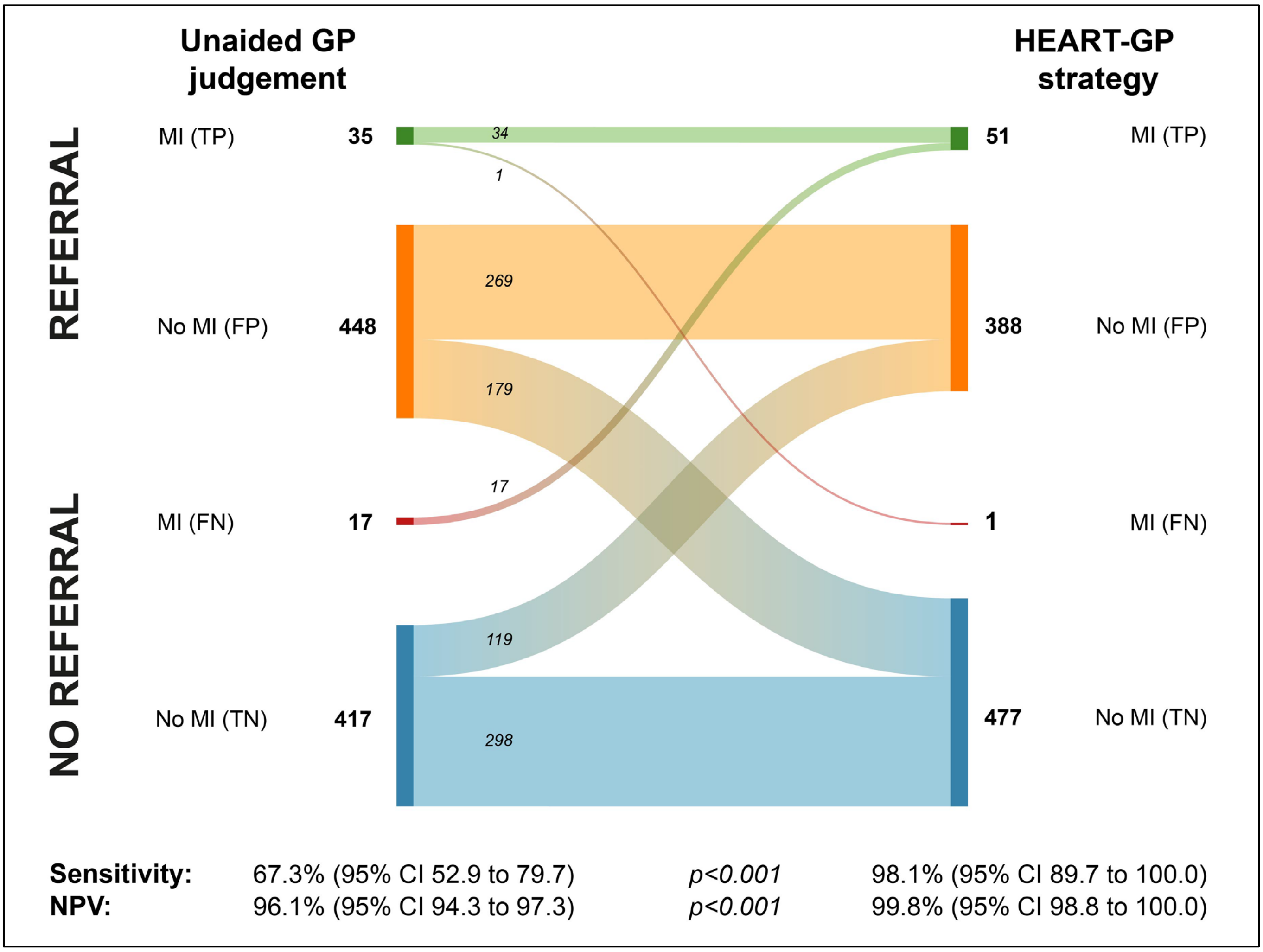


*FN: false-negative; FP: false-positive; GP: general practitioner; HEART-GP: the HEART-GP strategy; MI: myocardial infarction; NPV: negative predictive value; TN: true-negative; TP: true-positive; 95% CI: 95% confidence interval.*

*Reclassification of referral status by GPs after using the HEART-GP strategy (right column) compared with unaided judgement (before troponin test results were available; left column) for the secondary safety outcome (myocardial infarction). Sensitivity and NPV are provided for each strategy, with p-values for statistical difference between both strategies.*

**Supplementary Table S1. POCT errors and 6-week MACE.**

| POCT status | MACE | No MACE | Total |
|---|---|---|---|
| **POCT error/no result** | 7 | 33 | 40 |
| **No POCT error** | 55 | 822 | 877 |
| **Total** | 62 | 855 | 917 |

*MACE: major adverse cardiovascular events; POCT: point-of-care test.*

**Supplementary Table S2. MACE events classified as no urgent referral by HEART-GP.**

| | |
|---|---|
| **Case 1** | A man (age: 75-80 years) presented with 10 hours of chest pain. The POCT troponin concentration was 12.0 ng/L, and neither unaided GP assessment nor HEART-GP indicated urgent referral. During follow-up, cardiology assessment raised suspicion of unstable angina: PET-imaging showed anterior ischaemia, and coronary angiography demonstrated significant proximal LAD disease with additional RCA and RCX lesions. No urgent revascularisation was performed. |
| **Case 2** | A woman (age: 80-85 years) presented with chest pain lasting more than 24 hours. The patient was in palliative care for metastatic cholangiocarcinoma with limited life expectancy. The POCT troponin concentration was 8.3 ng/L. Both unaided GP assessment and HEART-GP indicated no urgent referral, with the treating GP explicitly mentioning the patient's metastatic disease as reason not to refer. The patient died within the follow-up window due to metastatic cholangiocarcinoma, and the event was adjudicated as non-cardiac death. |
| **Case 3** | A man (age: 70-75 years) with previous PCI presented with on and off chest pain that had been existing for 24 hours. At the OOH-PC, ECG was normal, POCT troponin was low at 2.2 ng/L, and symptoms resolved after nitroglycerin. HEART-GP did not indicate urgent referral. Later that evening, chest pain recurred and anterior STEMI was diagnosed, requiring primary PCI of the LAD. The event was adjudicated as unstable angina progressing to STEMI. |

*HEART-GP: the HEART-GP strategy; POCT: point-of-care test; STEMI: ST-elevation myocardial infarction.*
